# Multiplexable virus detection by CRISPR-Cas9-mediated strand displacement

**DOI:** 10.1101/2022.11.23.22282642

**Authors:** Rosa Márquez-Costa, Roser Montagud-Martínez, María-Carmen Marqués, María Heras-Hernández, Eliseo Albert, David Navarro, José-Antonio Daròs, Raúl Ruiz, Guillermo Rodrigo

**Affiliations:** Institute for Integrative Systems Biology (I2SysBio), CSIC – University of Valencia, 46980 Paterna, Spain; Microbiology Service, Clinic University Hospital, INCLIVA Biomedical Research Institute, 46010 Valencia, Spain; Department of Microbiology, School of Medicine, University of Valencia, 46010 Valencia, Spain; Instituto de Biología Molecular y Celular de Plantas (IBMCP), CSIC – Universitat Politècnica de València, 46022 Valencia, Spain

**Author notes:** Equal contribution to this work (Rosa Márquez-Costa, Roser Montagud-Martínez).

**Keywords:** CRISPR diagnostics, DNA nanotechnology, Synthetic biology

## Abstract

Recurrent disease outbreaks caused by different viruses, including the novel respiratory virus SARS-CoV-2, are challenging our society at a global scale; so better and handier virus detection methods would enable a faster response. Here, we present a novel nucleic acid detection strategy based on CRISPR-Cas9, whose mode of action relies on strand displacement rather than on collateral catalysis, using the *Streptococcus pyogenes* Cas9 nuclease. Given a pre-amplification process, a suitable molecular beacon interacts with the ternary CRISPR complex upon targeting to produce a fluorescent signal. We show that SARS-CoV-2 DNA amplicons generated from patient samples can be detected with CRISPR-Cas9. Moreover, we show that CRISPR-Cas9 allows the simultaneous detection of different DNA amplicons with the same nuclease, either to detect different SARS-CoV-2 regions or different respiratory viruses. Collectively, this CRISPR-Cas9 R-loop usage for molecular beacon opening (COLUMBO) platform allows a multiplexed detection in a single tube, complements the existing CRISPR-based methods, and displays diagnostic potential.

## INTRODUCTION

Infectious diseases defy the modern lifestyle of our societies, and it is increasingly evident that better and handier detection methods of viruses and bacteria would facilitate their control. Of note, coronavirus disease 2019 (COVID-19) pandemic caused by severe acute respiratory syndrome coronavirus 2 (SARS-CoV-2) [1] has highlighted the challenges in diagnostics of viral infections. A fast and confident diagnostic contributes to significantly reducing the transmission of the virus in the community and allows early therapeutic actions that can mitigate acute outcomes of infection. Currently, reverse transcription quantitative polymerase chain reaction (RT-qPCR) is the gold-standard diagnostic technique of infectious diseases in the clinic due to its high sensitivity and specificity [2]. However, when a rapid and massive intervention is required, such as in a pandemic context, alternative techniques that can bypass, at least in part, the need for expensive equipment and well-trained personnel are highly required [3].

Clustered regularly interspaced short palindromic repeats (CRISPR) systems are being repurposed in recent years for diagnostic applications [4, 5]. Owing to the ability of some CRISPR-associated (Cas) proteins to display a collateral catalytic activity upon target recognition, a sensitive and specific nucleic acid detection is possible. In combination with isothermal amplification techniques [6], sensitivities at the attomolar scale (*i.e*., about one copy per microliter) and specificities at one nucleotide resolution have been achieved, atop of bypassing the dependence on qPCR equipment. In this regard, the use of CRISPR systems may represent a suitable alternative for the diagnostics of infectious diseases in the clinic and also in the field. Notably, these systems have been already applied to detect SARS-CoV-2 in clinical samples [7]. Indeed, different assays based on CRISPR-Cas12 [7–9] or CRISPR-Cas13 [10–12] have been implemented for the detection of SARS-CoV-2 (even the direct detection of the virus without pre-amplification has been possible with the *Leptotrichia buccalis* Cas13a nuclease [12]).

However, the multiplexed detection remains challenging due to the non-specificity of that collateral catalytic activity. Certainly, sensing different elements at a time can be of utility for determining the presence of co-infecting pathogens or even for genotyping and identifying diverse mutations. To overcome this limitation, orthogonal Cas proteins can be employed to achieve multiplexed nucleic acid detection in a single reaction (*e.g*., by using a Cas12 to target DNA and a Cas13 to target RNA) [13]. Nevertheless, the number of nucleic acids that can be detected simultaneously is limited by the number of different Cas proteins involved in the assay. The use of droplets constitutes an alternative to distribute the detection by performing specific reactions in different compartments, thereby allowing a multiplexed detection with just one CRISPR system [14], in addition to descending the limit of detection [15]. Moreover, a recent development exploited the formation of non-canonical CRISPR RNAs for multiplexed RNA detection with Cas9 revealed by gel electrophoresis [16]. In any case, we still need to develop further methods easy to implement for multiplexed nucleic acid detection, especially to achieve point-of-care applications in emergency scenarios.

In this work, we present a novel nucleic acid detection approach based on CRISPR- Cas9 aimed at fulfilling the aforementioned gap. We exploited the absence of collateral catalytic activity of *Streptococcus pyogenes* Cas9 to develop a simple procedure based on CRISPR-mediated strand displacement [17] and fluorogenic molecular beacons [18] that allows a direct multiplexed detection of nucleic acids in a single tube. We called this platform COLUMBO (CRISPR-Cas9 R-loop usage for molecular beacon opening). First, we demonstrate that this can be applied to detect SARS-CoV-2. Second, we demonstrate a simultaneous detection of three different genomic regions of this coronavirus, as well as the potential application of detecting three different viruses in the sample or even discriminating SARS-CoV-2 variants without the need of sequencing.

## RESULTS

### Rational design of COLUMBO

COLUMBO requires a pre-amplification step to generate a suitable double-stranded DNA fragment from the nucleic acid of interest (DNA or RNA). This can be done by PCR or by an alternative method running isothermally, such as recombinase polymerase amplification (RPA) [19]. Importantly, the amplified DNA molecule needs to harbor a protospacer adjacent motif (PAM) for Cas9 binding (NGG). Then, the sequence-specific detection is accomplished thanks to interfacing a CRISPR- Cas9 reaction with an appropriately designed molecular beacon (single-stranded DNA, ssDNA) folding into a stem-loop structure. Once the R-loop is formed, the non-targeted DNA strand that has been displaced can interact with other nucleic acids supplied in *trans* [20]. Previously, we showed that this mechanism is instrumental to engineer toehold-free DNA circuits for logic computation [17]. Here, the interaction of the beacon with the displaced strand causes the reconfiguration of the former separating the fluorophore from the quencher, thereby producing a fluorescent signal (**Fig. 1a**).

**Figure 1:**
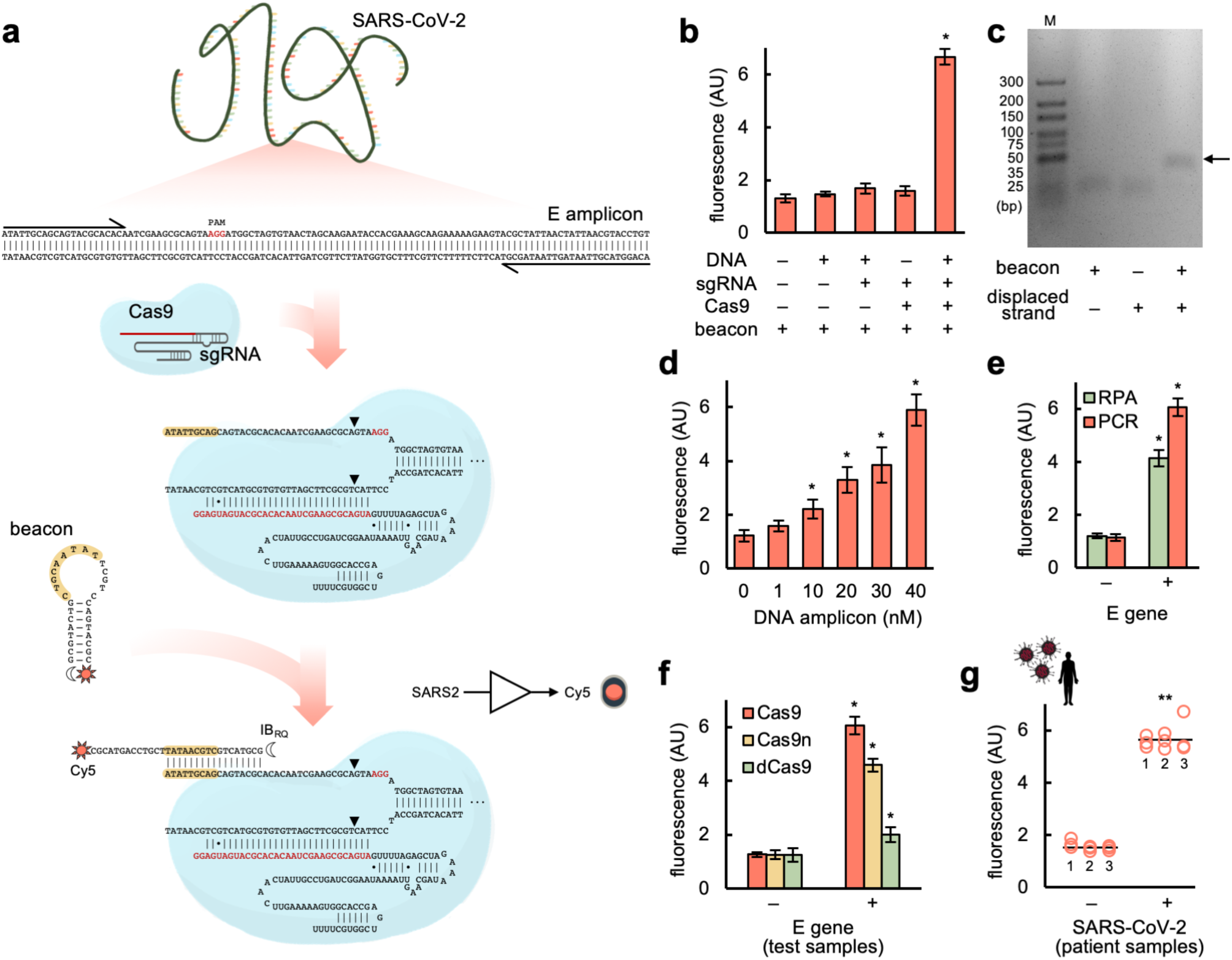
SARS-CoV-2 detection through a CRISPR-Cas9-based strand displacement reaction. a) Schematics of the global reaction of amplification and detection of a DNA product from SARS-CoV-2 E gene (PCR primers drawn at the ends), containing a PAM (shown in red) for Cas9 recognition. A preassembled CRISPR-Cas9 ribonucleoprotein targeting the amplicon (sgRNA spacer marked in red) was then able to displace a strand so that the molecular beacon could interact with and change its conformation (seed region for this interaction marked in yellow). The molecular beacon was labelled with the fluorophore Cy5 (sun icon) in the 3’ end and the dark quencher IB_RQ_ (moon icon) in the 5’ end. Wobble base pairs denoted by dots. b) Fluorescence-based characterization of the detection; amplifications performed by PCR. c) Gel electrophoretic assay to reveal the interaction between the molecular beacon and the displaced strand from the DNA amplicon (the arrow marks the intermolecular complex). M, molecular marker. d) Effect of the DNA amplicon concentration on the output fluorescence signal. e) Detection of the DNA amplicon generated by PCR or RPA (isothermal method). f) Effect of different versions of Cas9 (Cas9, Cas9n, or dCas9) on the output signal. g) Detection of SARS- CoV-2 in patient samples (6 patients, 3 reactions per patient); amplifications performed by RT-PCR. Error bars correspond to standard deviations (*n* = 3). *Statistical significance with test samples (Welch’s *t*-test, two-tailed *P* < 0.05). **Statistical significance with patient samples (Welch’s *t*-test, two-tailed *P* < 0.001).

We envisioned a system in which the PAM-distal region of the displaced strand is responsible for the interaction with the beacon. This interaction is seeded by the pairing of some nucleotides located in the loop of the beacon with the complementary nucleotides located in 5’ end of the displaced strand, ensuring a low activation energy barrier [21]. The beacon is not fully complementary to the displaced strand (only one-half binds to it). Moreover, to limit the potential interaction of the beacon with the single guide RNA (sgRNA), the spacer of the sgRNA excludes the seed region of the displaced strand. The spacer also harbors a mutation in the 5’ end region to form a wobble base pair with the targeted DNA strand [22]. The R-loop opens spontaneously at the PAM-distal region as a result of a low melting temperature, thereby exposing to the solvent the 5’ end of the displaced strand. Consequently, the beacon only opens when the ternary CRISPR complex (DNA-sgRNA-Cas9) is formed (**Fig. S1**).

### SARS-CoV-2 detection with COLUMBO

Using the Charité (Berlin) E-Sarbeco primers [23], we generated a suitable DNA amplicon for COLUMBO by PCR from a test sample based on the SARS-CoV-2 E gene. The amplified material was purified to remove elements potentially interfering with the molecular beacon. An sgRNA was designed to exploit a PAM located at an appropriate position (**Fig. 1a**), *in vitro* transcribed from a DNA template, and assembled with a Cas9 given from a commercial preparation. In turn, a molecular beacon appropriately designed was chemically synthesized, labelling its 3’ end with the fluorophore cyanine 5 (Cy5) and its 5’ end with the dark quencher Iowa Black RQ (IB_RQ_). Then, we added the sgRNA-Cas9 ribonucleoprotein to the reaction to target the amplified DNA (detection of the nucleic acid of interest) and the beacon to produce a red fluorescent signal upon interaction with the displaced strand in the PAM-distal region. Remarkably, COLUMBO displayed good performance, with a dynamic range of more than 3-fold change in red fluorescence and no apparent opening of the beacon in response to the DNA amplicon or the sgRNA alone (**Fig. 1b**). The ability of the beacon to interact with the displaced strand was also assessed by agarose gel electrophoresis (**Fig. 1c**). Furthermore, we performed a set of reactions with increasing concentrations of the DNA amplicon, observing proportionality between the input and output signals (**Fig. 1d**). Thus, COLUMBO might be used to quantify a given DNA in the sample ranging from the nanomolar scale.

Next, we tested the ability of using RPA instead of PCR to generate the DNA amplicon in combination with COLUMBO, as this is important to achieve point-of-care applications. Our results indicate that both methods are suitable, having used the very same primers (**Fig. 1e**). A slightly higher fluorescent signal in presence of the target DNA was produced after PCR, while the pre-amplification process was faster with RPA. In terms of sensitivity, 1 copy/µL in the sample was detected irrespective of the amplification method (**Fig. S2**). In addition, we inspected the impact of the catalytic activity of Cas9 on the performance of COLUMBO. To this end, we used three different versions of Cas9: the wild-type nuclease, the Cas9 H840A nickase (Cas9n), which only cleaves the non-targeted strand, and the catalytically dead Cas9 protein (dCas9), which does not produce any cleavage [24]. Both Cas9 and Cas9n produced a substantial fold change in fluorescence upon detection, although higher in the case of Cas9 (**Fig. 1f**). However, dCas9 failed in reaching such a performance, despite a significant differential readout was still possible. Arguably, the cleavage of the non-targeted strand confers more translational and rotational freedom to facilitate the interaction with the beacon [25]. We also found Cas9 and Cas9n to have equal activity on displacing that strand, but lower in the case of dCas9 (**Fig. S3**). Motivated by these results, we decided to apply COLUMBO to detect SARS-CoV-2 in patient samples. Nasopharyngeal swabs from people diagnosed as positive or negative in viral infection by RT-qPCR in the hospital were collected [26]. We reconfirmed the infections by RT-qPCR in our lab (**Fig. S4**). After RT-PCR amplification with the Charité E-Sarbeco primers (without RNA extraction), COLUMBO displayed marked differential readouts that were useful to discriminate the presence of the virus (**Fig. 1g**). These results demonstrate the potential suitability of COLUMBO to perform clinical diagnostics in a simple and effective way.

### Evaluation of modifications of COLUMBO

To avoid the purification step after the pre-amplification reaction, the region targeted by the molecular beacon should distinguish from those targeted by the primers and the sgRNA. In this regard, a strategy based on cleaving the resulting DNA amplicon in the PAM-distal region with a restriction enzyme was devised (**Fig. S5**). We achieved a successful detection with no apparent leakage following this approach (dynamic range of more than 2.5-fold in red fluorescence). In addition, we investigated the use of the PAM-proximal region of the displaced strand to interact with the molecular beacon. A new beacon targeting the N1 amplicon was designed. We found a significant opening of the beacon as a result of the interaction, but we also noticed an unwanted interaction with the sgRNA, which reduced the net dynamic range of the system (**Fig. S6**).

### Multiplexed SARS-CoV-2 detection with COLUMBO

We then moved forward to perform the simultaneous detection of different genomic regions of SARS-CoV-2. This is important to minimize the rate of false positives. The Centers for Disease Control and Prevention (CDC) N1 and N2 primers [23] were used together with the Charité E-Sarbeco primers to generate three different DNA amplicons by PCR from a test sample based on the SARS-CoV-2 N and E genes. Suitable PAMs were found in these amplicons (**Fig. 3a**). We designed the corresponding sgRNAs and molecular beacons according to the aforementioned specifications. The new beacons to detect the N1 and N2 amplicons were labelled with the fluorophores carboxyfluorescein (FAM) and carboxytetramethylrhodamine (TAMRA), respectively, in their 3’ end and with the dark quencher Iowa Black FQ (IB_FQ_) in their 5’ end. No significant interferences were observed when using simultaneously these three fluorophores (**Fig. S7**). Notably, COLUMBO allowed the precise detection of the different genomic regions having performed a series of combinatorial amplifications, with a minimal 3-fold change in relative fluorescence (**Fig. 2b**). In addition, we performed a multiplexed RT-PCR amplification over patient samples with all primers. COLUMBO gave a positive signal in the three fluorescence channels in the case of patients diagnosed as infected by SARS-CoV-2 in the hospital (**Fig. 2c**).

**Figure 2:**
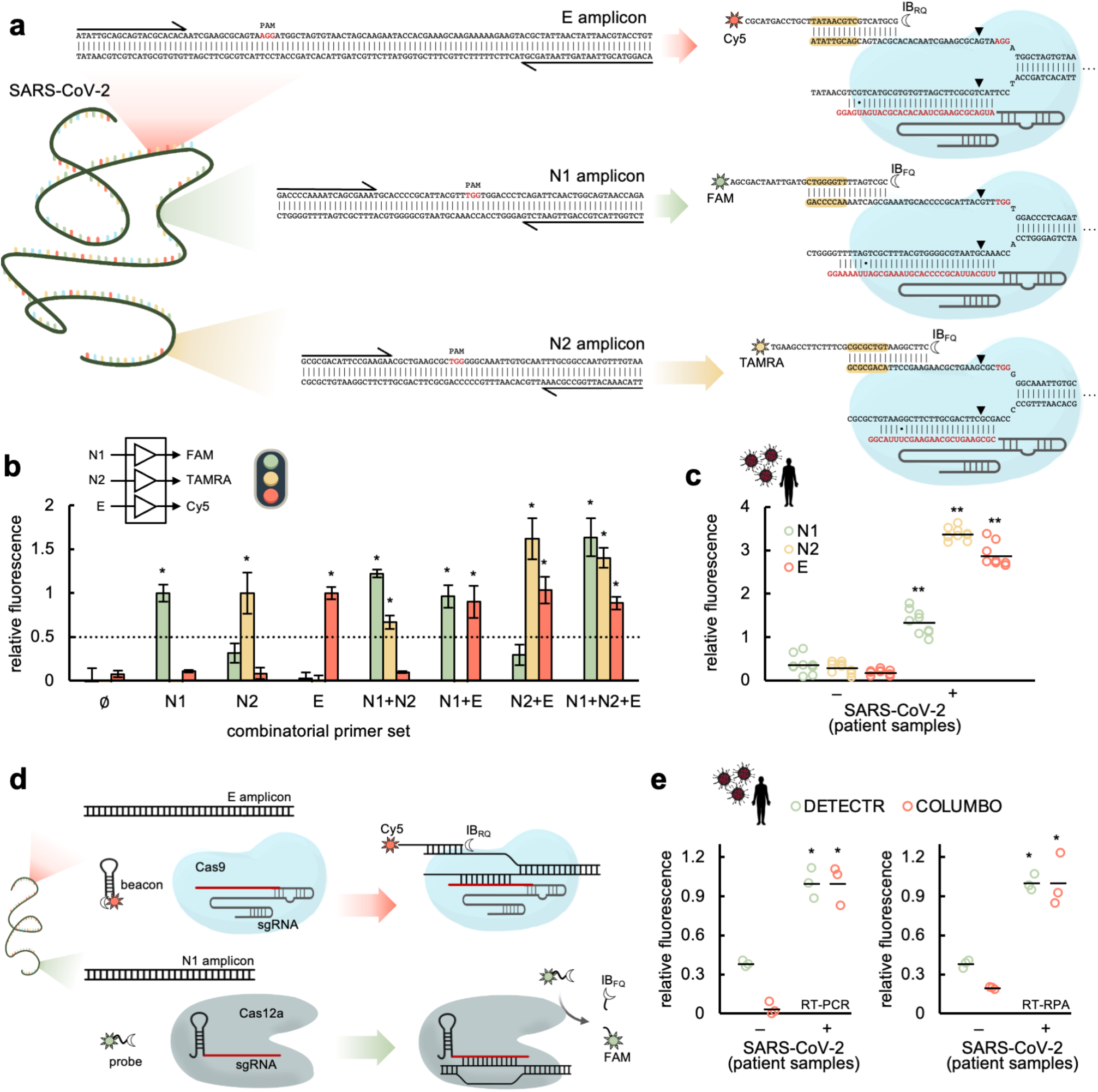
Multiplexed SARS-CoV-2 detection through CRISPR-Cas9-based strand displacement reactions. a) Schematics of the global reactions of amplification and detection of three different DNA products from SARS-CoV-2 (from E and N genes, PCR primers drawn at the ends), containing each a PAM (shown in red) for Cas9 recognition. Preassembled CRISPR-Cas9 ribonucleoproteins targeting the amplicons (sgRNA spacers marked in red) and appropriate molecular beacons were used for the detection (seed regions for the beacon-displaced strand interaction marked in yellow). The molecular beacons were labelled with Cy5 and IB_RQ_ (for E detection), FAM and IB_FQ_ (for N1 detection), and TAMRA and IB_FQ_ (for N2 detection). Wobble base pairs denoted by dots. b) Fluorescence-based characterization of the detection by performing amplifications with combinatorial sets of primers by PCR. The detection threshold (given by the dotted line) was set to 0.5, indicating that the difference in relative fluorescence was more than 2-fold. c) Multiplexed detection of SARS-CoV-2 in patient samples (6 patients, 3 reactions per patient); amplifications performed by RT-PCR. d) Schematics of the reaction of detection with COLUMBO and DETECTR of two different DNA products from SARS-CoV-2 (from E and N genes). A ssDNA probe labelled with FAM and IB_FQ_ was used for DETECTR (for N1 detection). e) Fluorescence-based characterization of the detection by performing multiplexed amplifications by RT-PCR or RT-RPA (2 patients, 3 reactions per patient). Error bars correspond to standard deviations (*n* = 3). *Statistical significance with test samples (Welch’s *t*-test, two-tailed *P* < 0.05, and relative fluorescence > 0.5). **/*Statistical significance with patient samples (Welch’s *t*-test, two-tailed *P* < 0.001/0.05).

**Figure 3:**
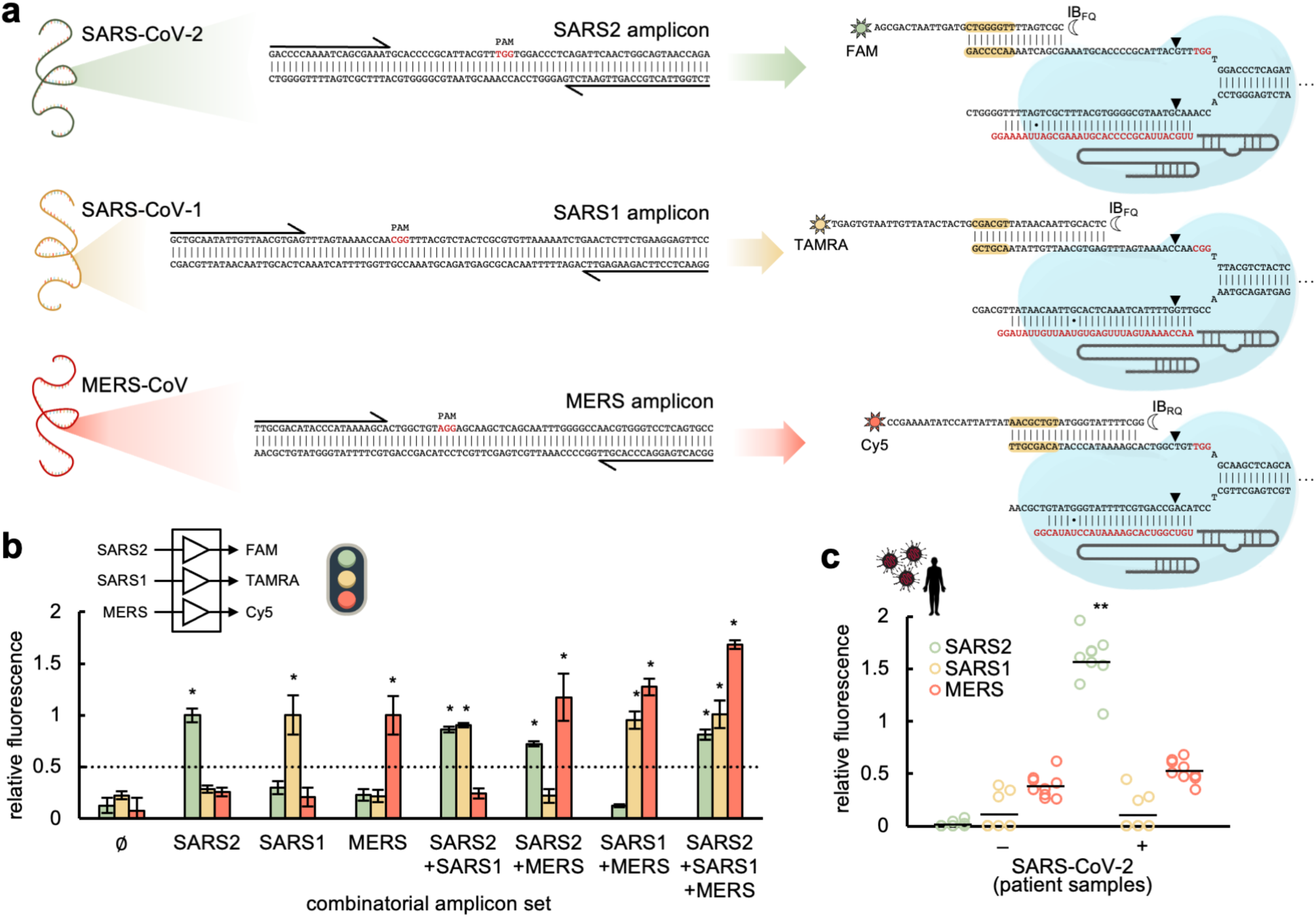
Multiplexed coronavirus detection through CRISPR-Cas9-based strand displacement reactions. a) Schematics of the global reactions of amplification and detection of DNA products from SARS-CoV-2, SARS-CoV-1, and MERS-CoV (PCR primers drawn at the ends), containing each a PAM (shown in red) for Cas9 recognition. Preassembled CRISPR-Cas9 ribonucleoproteins targeting the amplicons (sgRNA spacers marked in red) and appropriate molecular beacons were used for the detection (seed regions for the beacon-displaced strand interaction marked in yellow). The molecular beacons were labelled with FAM and IB_FQ_ (for SARS-CoV-2 detection), TAMRA and IB_FQ_ (for SARS-CoV-1 detection), and Cy5 and IB_RQ_ (for MERS-CoV detection). Wobble base pairs denoted by dots. b) Fluorescence-based characterization of the detection by working with combinatorial sets of DNA amplicons. The detection threshold (given by the dotted line) was set to 0.5, indicating that the difference in relative fluorescence was more than 2- fold. c) Differential detection of SARS-CoV-2 in patient samples (6 patients, 3 reactions per patient); amplifications performed by RT-PCR. Error bars correspond to standard deviations (*n* = 3). *Statistical significance with test samples (Welch’s *t*-test, two-tailed *P* < 0.05, and relative fluorescence > 0.5). **Statistical significance with patient samples (Welch’s *t*-test, two-tailed *P* < 0.001).

Furthermore, we investigated the possibility of using DETECTR (DNA endonuclease-targeted CRISPR *trans* reporter) [5] in combination with COLUMBO to perform multiplexed detections. The structure of the molecular beacon together with a low concentration regime for Cas12a would prevent a premature degradation of the beacon as a result of the collateral catalytic activity of this nuclease upon targeting. Alternatively, a molecular beacon of RNA could be used. We focused on detecting the DNA amplicon generated with the CDC N1 primers with DETECTR, noting that it contains a PAM for Cas12a binding (TTTV) [7], and the amplicon generated with the Charité E-Sarbeco primers with COLUMBO (**Fig. 2d**). A suitable sgRNA was *in vitro* transcribed and assembled with the *Acidaminococcus sp.* Cas12a for DETECTR. After multiplexed RT-PCR or RT-RPA amplification over patient samples, we found that COLUMBO and DETECTR can work together to detect different SARS-CoV-2 genes (**Fig. 2e**).

### Differential virus detection with COLUMBO

Next, we assessed the ability of COLUMBO to detect simultaneously different viruses in the sample. Such a multiplexed detection is important because it may allow performing in the future differential diagnostics and uncovering mixed infections. We focused on three different coronaviruses: SARS-CoV-2, SARS-CoV-1, and Middle East respiratory syndrome coronavirus (MERS-CoV) [27]. The CDC N1 primers were considered for SARS-CoV-2, new primers were designed for SARS-CoV-1, and previously designed primers were taken for MERS-CoV [28]. We checked that each pair of primers only aligns with the cognate genome. Suitable PAMs were found in the corresponding amplicons (**Fig. 3a**). We then designed new sgRNAs and beacons to detect SARS-CoV-1 (signal from fluorophore TAMRA) and MERS-CoV (signal from fluorophore Cy5). The low GC content of the protospacer regions in these cases forced to design beacons with larger stems, in order to ensure intra- and intermolecular stability. Notably, COLUMBO allowed the precise detection of the different DNA amplicons added in a combinatorial way, with a minimal 2.4-fold change in relative fluorescence as before (**Fig. 3b**). In addition, we performed a multiplexed RT-PCR amplification over patient samples with all primers. COLUMBO gave a positive signal only in the green fluorescence channel, corresponding to SARS-CoV-2, in the case of patients diagnosed as infected in the hospital (**Fig. 3c**). These results indicate that COLUMBO is a suitable method to achieve a direct multiplexed detection of nucleic acids with no need of gel electrophoresis, in contrast to previous work [16].

Finally, we tested if COLUMBO, in addition to detect SARS-CoV-2, can reveal specific mutations. This would be important to provide a cost-effective genetic perspective about the transmission dynamics in the pandemic [29]. We hypothesized that it could be possible to perform a multiplexed detection with two sgRNAs, one targeting a conserved region to confirm the infection by SARS-CoV-2 and another targeting a mutable region to identify a specific viral genotype (**Fig. 4a**). Mutations in the PAM would compromise the binding of the Cas protein, while mutations in the protospacer the binding of the beacon (in the PAM-distal region) and the sgRNA (in the PAM-proximal region; **Fig. S8**). As a proof of concept, we here focused on the H69-V70 deletion of six nucleotides in the S gene (S- delH69-V70) identified in the Alpha variant [30]. Using test samples containing the E and S/S-delH69-V70 amplicons, the intended multiplexed detection was successfully accomplished, indicating that the corresponding sgRNA does not interact with the wild-type S amplicon (**Fig. 4b**). We also considered a deletion of three nucleotides in the N gene (N-delQ9) identified in a Delta variant sublineage [31], which we were able to detect in a multiplexed fashion using test samples containing the E and N1/N1-delQ9 amplicons (**Fig. S9**). These examples suggest that, provided suitable sgRNAs and beacons are designed, COLUMBO could be applied to identify further SARS-CoV-2 mutants to limit the use of sequencing.

**Figure 4:**
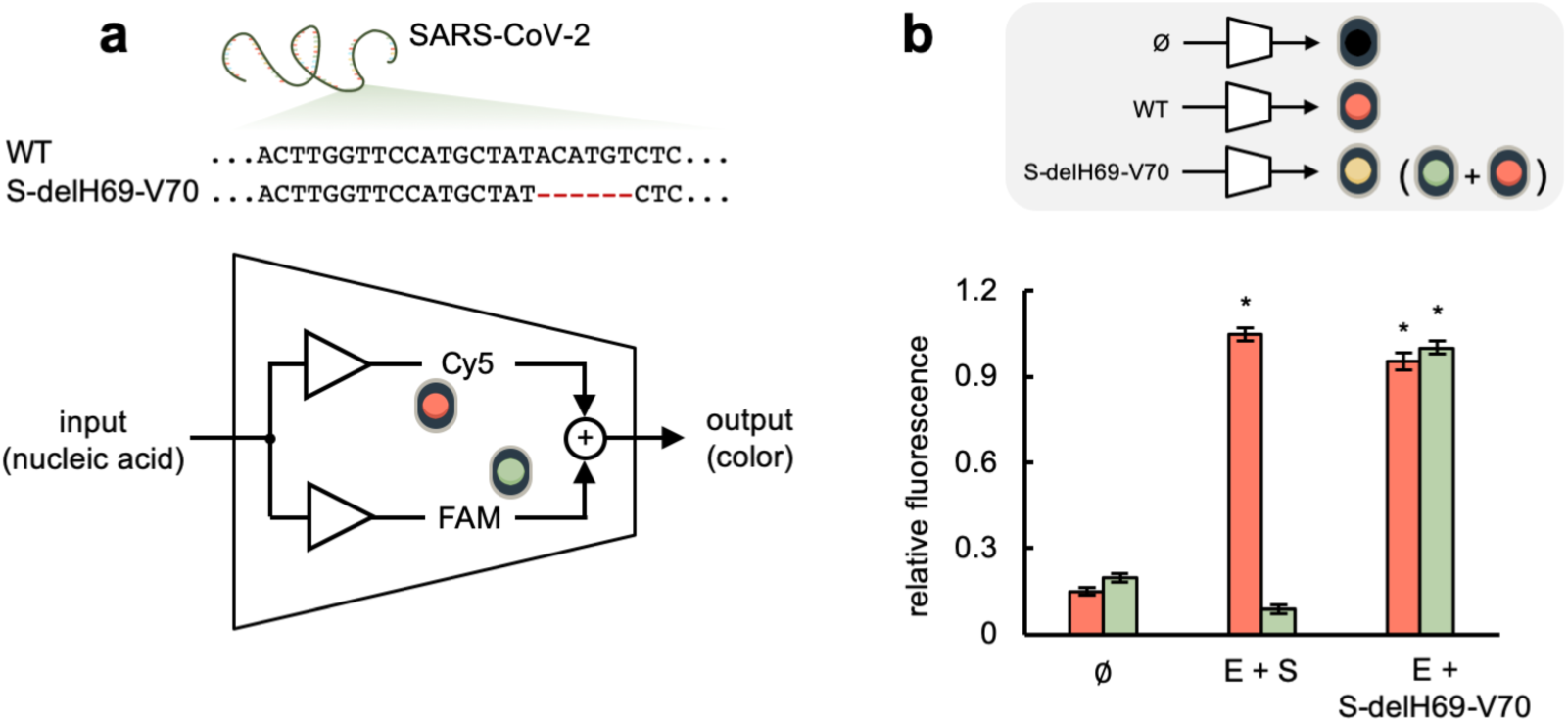
Mutant SARS-CoV-2 detection through CRISPR-Cas9-based strand displacement reactions. a) Schematics of an electronic circuit implementing a molecular program for detection: if the sample is free of SARS-CoV-2, there is no light signal; if it contains the wild-type SARS-CoV-2, a red signal is obtained (only from Cy5); if it contains a SARS-CoV-2 that carries the mutation S-delH69-V70, red and green signals are obtained (from Cy5 and FAM), *i.e.*, a “yellow” signal. b) Fluorescence-based characterization of the detection by working directly with DNA amplicons: none, E and S (simulating the wild-type SARS-CoV-2), and E and S-delH69-V70 (simulating a mutant SARS-CoV-2, Alpha variant). Error bars correspond to standard deviations (*n* = 3). *Statistical significance with test samples (Welch’s *t*-test, two-tailed *P* < 0.05).

## DISCUSSION

We have enlarged the CRISPR-based detection toolkit with the CRISPR-Cas9 system on which COLUMBO relies. As in the case of SHERLOCK (specific high-sensitivity enzymatic reporter unlocking) [4] and DETECTR [5], a pre-amplification process is required. However, COLUMBO is based on strand displacement (hybridization) and not on a collateral catalytic activity. That is, the sgRNA-Cas9 ribonucleoprotein gives the specificity of the detection by targeting the resulting DNA amplicon, and the interaction between the displaced DNA strand and a molecular beacon gives the fluorescence readout. Consequently, different DNA amplicons can be detected with the same nuclease (Cas9) and without the need of complex setups to run parallel microreactions [14, 15], which is an advance in terms of broad usability and standardization. Remarkably, we have applied this novel approach with success to detect SARS-CoV-2 in patient samples, thereby envisioning a diagnostic potential. We also showed that COLUMBO and DETECTR can work together, a feature that is important to boost intricacy in CRISPR diagnostics.

In our designer scheme, the native Cas9 from *S. pyogenes* is exploited, but the use of engineered versions of Cas9 with enhanced specificity [32] could make the detection of virus variants with subtle specific mutations more plausible. COLUMBO might be run after RT-qPCR completion as a simple virus genotyping step in order to complement the quantitative detection in the clinic with relevant information for patient prognosis and epidemiological surveillance. Further work should also refine the approach to make it even more streamlined (*i.e*., joining amplification and detection). On the other hand, increasing developments on portable fluorescence microscopy coupled to mobile phones, already applied to detect SARS-CoV-2 [12, 33], are appealing and might be used to characterize our CRISPR-Cas9 reactions. A cell-free expression system might also be interfaced to achieve a detection with a colorimetric readout [34], given that strand displacement allows nucleic acid conversion [17]. This would enable point-of-care diagnostics and field-deployable testing.

Finally, we envision the application of computational methods to automate the sequence design of the different nucleic acid species that COLUMBO requires, given a series of energetic and structural specifications [35, 36]. This would allow ending with species with suitable intra- and intermolecular stability, while minimizing undesired cross-interactions (*e.g*., between the molecular beacon and the sgRNA). One limitation of COLUMBO is the sometimes-moderate dynamic range of the fluorescence signal, which may difficult an accurate detection of the virus, especially in the case of multiplexing. In this regard, the combination of computational sequence design with systematic functional screening might allow optimizing the system. In addition, computational methods might contribute to the engineering of more complex nucleic acid circuits not only to detect a specific sequence but to perform a logic computation from multiple input signals (*e.g*., to inform about the presence of two or more viruses in the sample with just one fluorophore) [37]. Within this extended diagnostic framework, the activity of the sgRNAs might be conditional to the presence or absence of further strands to have an additional layer of operation [38]. All in all, due to its specificity, multiplexing capability, compatibility, and easy usage, we expect COLUMBO to provide exciting prospects in the field of viral diagnostics and DNA/RNA-based computation.

## METHODS

### Test samples

For single detection, a test sample was generated with a plasmid containing the SARS-CoV-2 E gene (IDT) at 10^3^ copies/µL (different dilutions down to 1 copy/µL were also made). Additional test samples were generated for multiplexed detection. First, a test sample was generated by mixing two plasmids containing the SARS-CoV-2 N and E genes (IDT) each at 10^3^ copies/µL. Second, different combinatorial samples of three DNA amplicons from different viruses were prepared. The DNA amplicon from SARS-CoV-2 was generated by PCR from the aforementioned plasmid with the CDC N1 primers, and the DNA amplicons from SARS-CoV-1 and MERS-CoV were chemically synthesized (IDT). Third, two DNA amplicons from the S gene, one being the wild-type version and another carrying the amino acid H69-V70 deletion [30], and a DNA amplicon from the N gene carrying the CAG deletion at position 28298 (deleting the ninth residue Q) [31] were chemically synthesized.

### Patient samples

Nasopharyngeal swabs corresponding to infected and non-infected patients with SARS-CoV-2 (RT-qPCR diagnostics) were obtained from the Clinic University Hospital of Valencia (Spain). Samples were inactivated by heat shock (30 min at 60 °C) before proceeding. No RNA extraction was performed. The ethical committee of the Clinic University Hospital approved this study (order #2020/221).

### Primers

The CDC N1 and N2 primers were used to amplify two different N gene regions from SARS-CoV-2, and the Charité E-Sarbeco primers were used to amplify one E gene region [23]. The Charité E-Sarbeco primers were used for both PCR and RPA, while the CDC N1 primers were only used for PCR. Longer primers targeting the N1 region were designed for RPA. Primers to amplify the S gene were also designed here. In addition, a genomic region from SARS-CoV-1 could be amplified with newly designed primers, and a region from MERS-CoV could be amplified with the previously designed MERS-related N2 primers [28]. Sequences provided in **Dataset S1**.

### CRISPR elements

Three versions of *S. pyogenes* Cas9 were used (from IDT): the wild-type nuclease (Cas9), the Cas9 H840A nickase (Cas9n), and the catalytically dead Cas9 protein (dCas9) [24]. *Acidaminococcus sp.* Cas12a (from IDT) was also used to implement DETECTR reactions. In addition, sgRNAs were generated by *in vitro* transcription with the TranscriptAid T7 high yield transcription kit (Thermo) from DNA templates. sgRNAs were then purified by using the RNA clean and concentrator column (Zymo) and quantified in a NanoDrop. Sequences provided in **Dataset S1**.

### Molecular beacons

Different DNA oligonucleotides folding into a stem-loop structure and appropriately labelled were designed to hybridize with the displaced DNA strands from the CRISPR reactions. These probes were designed to have a seed region in the loop and of high GC content, as well as a melting temperature higher than 50 °C (**Fig. S10** shows a computational study on the size of the seed region). The correct folding and hybridization ability (with the target DNA, but not with the sgRNA) were checked with NUPACK [36]. Molecular beacons targeting the PAM-distal region were labelled in their 5’ end with a dark quencher (lBFQ or lBRQ) and in their 3’ end with a fluorophore (FAM, TAMRA, or Cy5). When targeting the PAM-proximal region, the beacon was labelled in its 5’ end with FAM and its 3’ end with lBFQ or Black Hole Quencher 1 (BHQ1). To ensure appropriate folding, molecular beacons were heated at 95 °C for 2 min and then cooled slowly to 25 °C prior to their use in the CRISPR reactions. Sequences provided in **Dataset S1**.

### Nucleic acid amplification by PCR

With test samples, 250 nM of forward and reverse primers, 200 µM dNTPs (NZYTech), 0.02 U/μL Phusion high-fidelity DNA polymerase (Thermo), 1x Phusion buffer, and 2 µL of sample were mixed for a total volume of 20 µL (adjusted with RNase-free water). The protocol was 98 °C for 30 s for denaturation, followed by 35 cycles of 98 °C for 10 s, 62 °C for 10 s, and 72 °C for 5 s for amplification. With patient samples, the TaqPath 1-step RT-qPCR master mix, CG (Applied) was used with 250 nM of forward and reverse primers and 4 µL of sample. The protocol was 50 °C for 15 min for RT, then 95 °C for 2 min for denaturation, followed by 35 cycles of 95 °C for 15 s and 62 °C for 60 s for amplification. In the case of multiplexed amplifications, each primer pair was also used at 250 nM. Reactions were incubated in a thermocycler (Eppendorf). PCR products were purified by using a DNA clean and concentrator column (Zymo) by centrifugation.

### Nucleic acid amplification by RPA

The TwistAmp basic kit (TwistDX) was used. With test samples, 480 nM of forward and reverse primers was added to 29.5 µL of rehydration buffer for a total volume of 45.4 µL (adjusted with RNase-free water). With patient samples, 500 U RevertAid (Thermo) and 50 U RNase inhibitor (Thermo) were added to the mix. In the case of multiplexed amplifications, each primer pair was used at 240 nM. The TwistAmp basic reaction pellet was resuspended with the resulting volume, then adding 2 µL of sample. To start the reaction, 280 mM magnesium acetate was added. In the case of multiplexed amplifications, each primer pair was also used at 240 nM. Reactions were incubated at 42 °C for 30 min in a thermomixer (Eppendorf), shaking 10 s at 300 rpm every 2 min. RPA products were purified by using the DNA clean and concentrator column by centrifugation.

### CRISPR-Cas9-based detection

CRISPR reactions were performed in 1x TAE buffer pH 8.5 (Invitrogen), 0.05% Tween 20 (Merck), and 12.5 mM MgCl_2_ (Merck) at a final volume of 20 µL. The CRISPR-Cas9 ribonucleoprotein, previously assembled at room temperature for 30 min, was added at 100 nM. In the case of test samples, 40 nM of amplified DNA (otherwise specified) was used per reaction. In the case of patient samples, 2 or 6 µL of amplified product was used per reaction for single or multiplexed detection. For the limit of detection assays, 2 µL of purified PCR product was used (5 µL in the case of RPA). Reactions were incubated at 37 °C for 20 min in a thermomixer (Eppendorf). The molecular beacon (beacons) was (were) added afterwards at 100 nM, followed by 5 min incubation at 37 °C. The beacons could also be added at the beginning of the CRISPR reaction to simplify the approach, obtaining similar results (**Fig. S5b**). For the multiplexed detection of the E and S-delH69-V70 amplicons, 200 nM of ribonucleoprotein targeting the S-delH69-V70 amplicon was used and the two beacons were added at the beginning.

### Combined CRISPR-Cas9- and CRISPR-Cas12a-based detection

CRISPR reactions were performed in 1x TAE buffer pH 8.5 (Invitrogen), 0.05% Tween 20 (Merck), and 12.5 mM MgCl_2_ (Merck) at a final volume of 20 µL. The CRISPR-Cas9 and CRISPR-Cas12a ribonucleoproteins, previously assembled at room temperature for 30 min, were added at 100 nM and 1.33 nM, respectively. The ssDNA probe for CRISPR-Cas12a (TTATT, labelled in its 5’ end with the fluorophore FAM and in its 3’ end with the dark quencher lBFQ) was also added at 100 nM. From patient samples, 4 µL of amplified product was used per reaction for multiplexed detection. Reactions were incubated at 37 °C for 20 min in a thermomixer (Eppendorf). The molecular beacon was added afterwards at 100 nM, followed by 5 min incubation at 37 °C.

### RT-qPCR

The TaqPath 1-step RT-qPCR master mix, CG was used. 2 µL of sample was mixed with 500 nM of forward and reverse primers (CDC N1), 250 nM of ssDNA probe (provided by IDT to detect the N gene), and 5 µL of the master mix for a total volume of 20 µL (adjusted with RNase-free water) in a fast microplate (Applied). Reactions were performed in a QuantStudio 3 equipment (Thermo) with this protocol: incubation at 25 °C for 2 min for uracil-N glycosylation, followed by 50 °C for 15 min for RT, followed by an inactivation step at 90 °C for 2 min, then followed by 40 cycles of amplification at 90 °C for 3 s and 60 °C for 30 s.

### Gel electrophoresis

Nucleic acid amplification (from plasmid or viral genome) was confirmed by gel electrophoresis (see examples in **Fig. S11**). For that, 2 µL of amplified product was used. Gel electrophoresis was also used to confirm the interaction between the molecular beacon and the displaced strand from the DNA amplicon (ssDNA molecule). For that, the nucleic acid species were introduced at 7.5 µM each in 20 µL of the CRISPR reaction buffer and were incubated for 30 min at room temperature. Samples were loaded on a 3% agarose gel prepared with 0.5x TBE buffer, which was run for 45 min at room temperature (110 V). Gels were stained using RealSafe (Durviz). The GeneRuler ultra-low range DNA ladder (10-300 bp, Thermo) was used as a marker.

### Fluorometry

Reaction volumes were loaded in a black 384-well microplate with clear bottom (Falcon), which was then placed in a fluorometer (Varioskan Lux, Thermo) to measure green, orange, and red fluorescence (measurement time of 100 ms, automatic range, and top optics). For FAM, excitation was at 495/12 nm and emission at 520/12 nm (green); for TAMRA, excitation was at 557/12 nm and emission at 583/12 nm (orange); and for Cy5, excitation was at 645/12 nm and emission at 670/12 nm (red). Fluorescence values were represented as absolute or relative. For the latter, the fluorescence values of the closed-form beacons were subtracted to correct the signals, which were then normalized by appropriate reference values. In the case of multiplexed reactions for the simultaneous detection of different SARS-CoV-2 genes or different coronaviruses, the normalization was with respect to the positive case where only one amplicon is present. In the case of reactions for mutant SARS-CoV-2 detection or combining Cas9 and Cas12a, the normalization was with respect to the positive case where all amplicons are present.

## Data Availability

All data produced in the present work are contained in the manuscript

## ACKNOWLEDGEMENTS

This work was supported by the Fondo Supera Covid-19 from CRUE and Banco Santander (grant COV-CRISPIS to GR), the CSIC PTI Salud Global (grant SGL2021-03-040 to GR) through the NextGenerationEU Fund (regulation 2020/2094), the Spanish Ministry of Science and Innovation (grants PGC2018-101410-B-I00 to GR and PID2020- 114691RB-I00 to JAD, both co-financed by the European Regional Development Fund), and the Regional Government of Valencia (grants SEJI/2020/011 and GVA- COVID19/2021/036 to GR). RMC was supported by a predoctoral fellowship from the Spanish Ministry of Science and Innovation (PRE2019-088531).

## COMPETING INTEREST

RMC, RMM, MHH, RR, and GR declare that are patenting the detection method presented in this work.

## SUPPLEMENTARY METHODS

### Note on the design of COLUMBO elements

In this work, we decided to use standard primers, such as the Charité E-Sarbeco primers and the CDC N1 and N2 primers for SARS-CoV-2. The occurrence of a PAM sequence for Cas9 (NGG) is a minimal requirement that can be met in many cases, as it occurs in the E and N gene amplicons. Nothing prevents using other primers. The position of the PAM sequence within the DNA amplicon determines the length of the sgRNA spacer. In principle, spacers of 20-40 nt should be adequate.

The precise sequence of the displaced strand conditions the sequence and secondary structure of the beacon. If it is rich in GC, the beacon can be designed with a shorter stem. The dynamic range of the beacon is uncertain *a priori*, so functional screening could be performed. Furthermore, for the beacon to interact efficiently with the displaced strand, the R-loop needs to be open in the PAM-distal end. Otherwise, the displaced strand has not sufficient freedom.

### Note on the application by RPA

In our nucleic acid amplifications by RPA, a shaking of 10 s at 300 rpm was applied every 2 min during the 30 min of the reaction. In principle, shaking is not required for RPA to work. However, in our hands, we found better amplifications with shaking, especially in the case of clinical samples. In some cases, RPA reactions without shaking produced no observable bands when revealed in a gel.

### Activity assessment of different Cas9 versions

A suitable dsDNA molecule, whose sequence is GGCTAAAGAGGAAGAGGACATGGTGAATTCGTAACT, was labelled with a fluorophore (FAM, in 5’) and a quencher (Iowa Black FQ, in 3’) in the PAM-distal ends. A suitable sgRNA, whose spacer is GGCUAAAGAGGAAGAGGACA, was used to perform the CRISPR reactions. These were done in 1x TAE buffer pH 8.5 (Invitrogen), 0.05% Tween 20 (Merck), and 12.5 mM MgCl_2_ (Merck) at a final volume of 20 µL. The sgRNA- Cas9 ribonucleoprotein was added at 100 nM and the labelled dsDNA at 20 nM. Here, Cas9, Cas9n, and dCas9 were used. Reactions were incubated at 37 °C for 20 min in a thermomixer (Eppendorf).

### Single-point mutation detection with CRISPR-Cas9

Different E gene amplicon variants harboring substitution mutations in the PAM or the protospacer (in the seed region, *i.e*., the PAM-proximal region) were chemically synthesized (IDT). Wild-type Cas9 and HiFi Cas9 from IDT were used. According to IDT, HiFi Cas9 has similar on-target potency to wild-type Cas9, but with significantly reduced off-target effects, then allowing for precise targeting. CRISPR reactions were performed as already indicated.

### Note on coupling the amplification and detection reactions

Because the beacon interacts in the PAM-distal region and the R-loop needs to open at that point, the forward primer (with respect to the PAM) overlaps with the region targeted by the beacon. Therefore, the primer may interfere in the detection reaction. This was solved by purifying the amplified material.

To avoid the purification step, we devised a strategy based on cleaving the resulting DNA amplicon in the PAM-distal region with a restriction enzyme. In this way, the forward primer (with respect to the PAM) and the beacon do not significantly overlap. A particular restriction site can be introduced with the primer or be present in the original sequence. In principle, different restriction enzymes may be used. Here, we used XbaI, which has good efficiency in our reaction buffer. We designed a new primer to amplify the SARS-CoV-2 E gene.

### CRISPR-Cas9-based detection with no prior purification

From test samples containing the SARS-CoV-2 E gene, DNA amplicons were generated by PCR as already indicated, this time with 45 cycles. Then, 6 µL of non-purified sample was used in the CRISPR reaction. This was performed in 1x TAE buffer pH 8.5 (Invitrogen), 0.05% Tween 20 (Merck), and 12.5 mM MgCl_2_ (Merck), with the sgRNA-Cas9 ribonucleoprotein at 100 nM, the molecular beacon at 100 nM, and XbaI at 0.5 U/µL, for a final volume of 20 µL. Reactions were incubated at 37 °C for 20 min in a thermomixer (Eppendorf).

## SUPPLEMENTARY FIGURES

**Figure S1:**
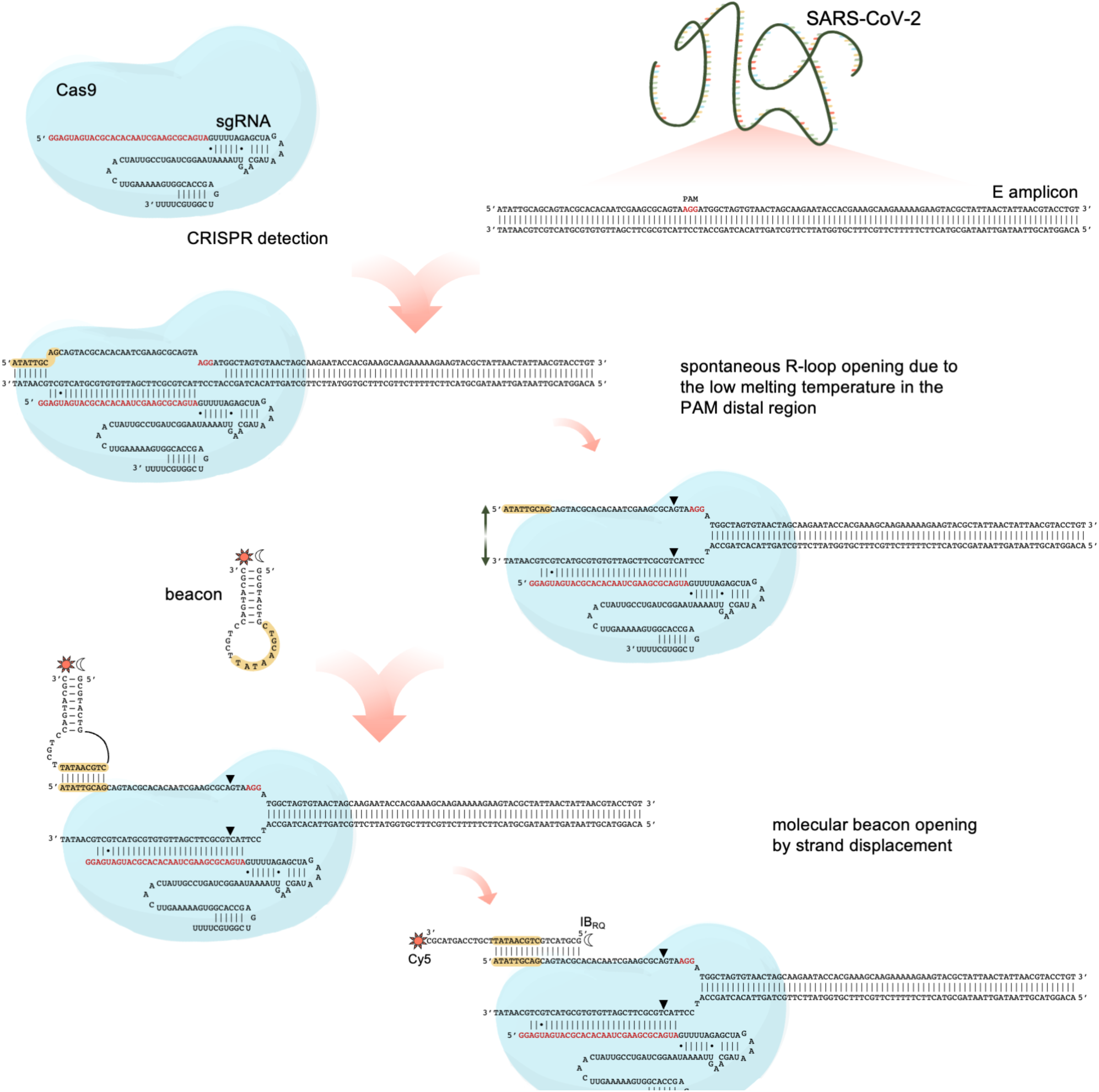
Detailed schematics of nucleic acid detection through CRISPR-Cas9- based strand displacement. In this example, a DNA amplicon from SARS-CoV-2 E gene was generated, containing a PAM (shown in red) for Cas9 recognition. A preassembled CRISPR-Cas9 ribonucleoprotein targeting the amplicon (sgRNA spacer marked in red) could then be used for sequence specific detection. The spacer starts by GG as it is *in vitro* transcribed by the T7 polymerase. The resulting R-loop was open by the PAM-distal region due to a low melting temperature in the DNA end (in this example, 7 base pairs). The displaced strand could interact with a properly designed molecular beacon (seed regions marked in yellow), which opens afterwards. The molecular beacon was labelled with the fluorophore Cy5 (sun icon) in the 3’ end and the dark quencher Iowa Black RQ (moon icon) in the 5’ end.

**Figure S2:**
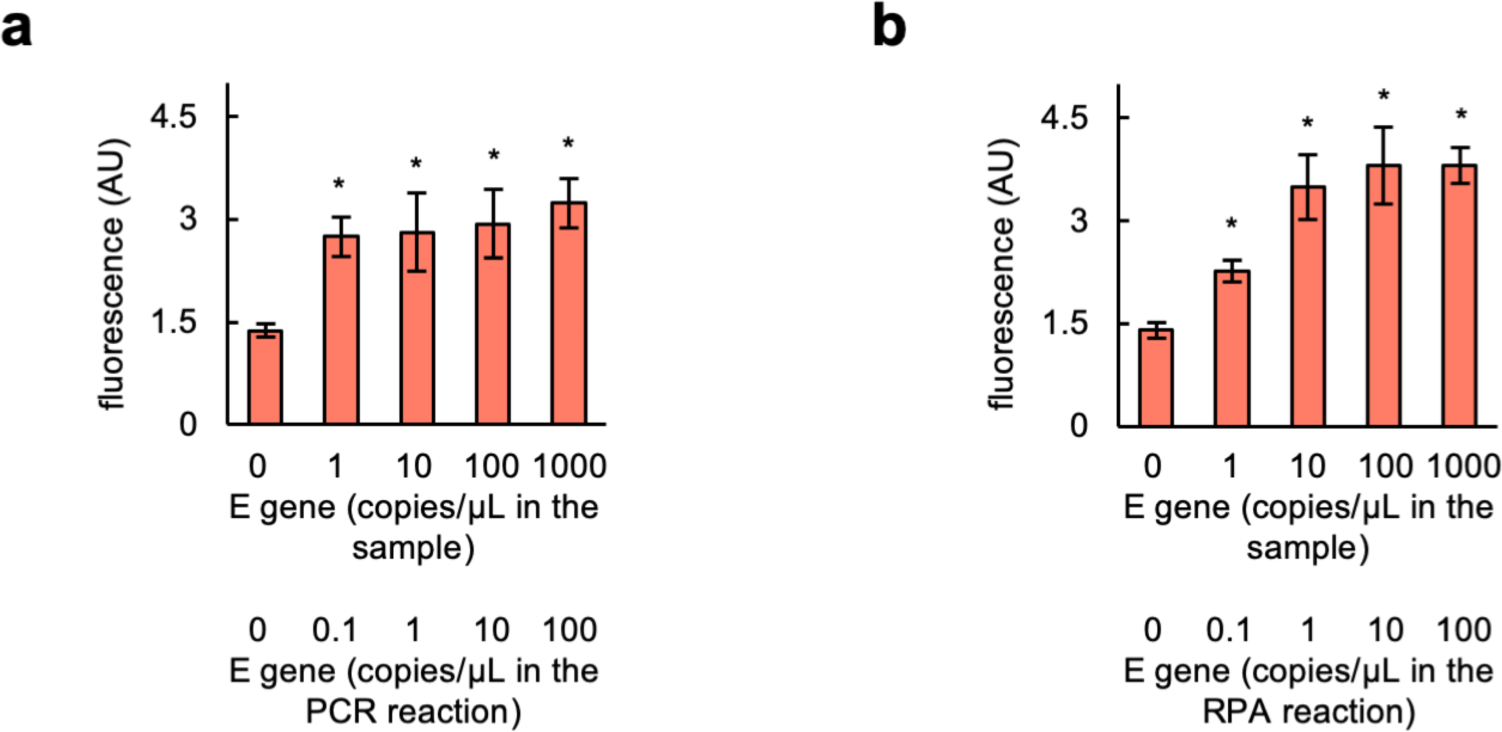
Limit of detection assays. a) Fluorescence-based characterization of the detection with COLUMBO-PCR. b) Fluorescence-based characterization of the detection with COLUMBO-RPA. Test samples containing the SARS-CoV-2 E gene at different concentrations were used. Error bars correspond to standard deviations (*n* = 3). *Statistical significance (Welch’s *t*-test, two-tailed *P* < 0.05).

**Figure S3:**
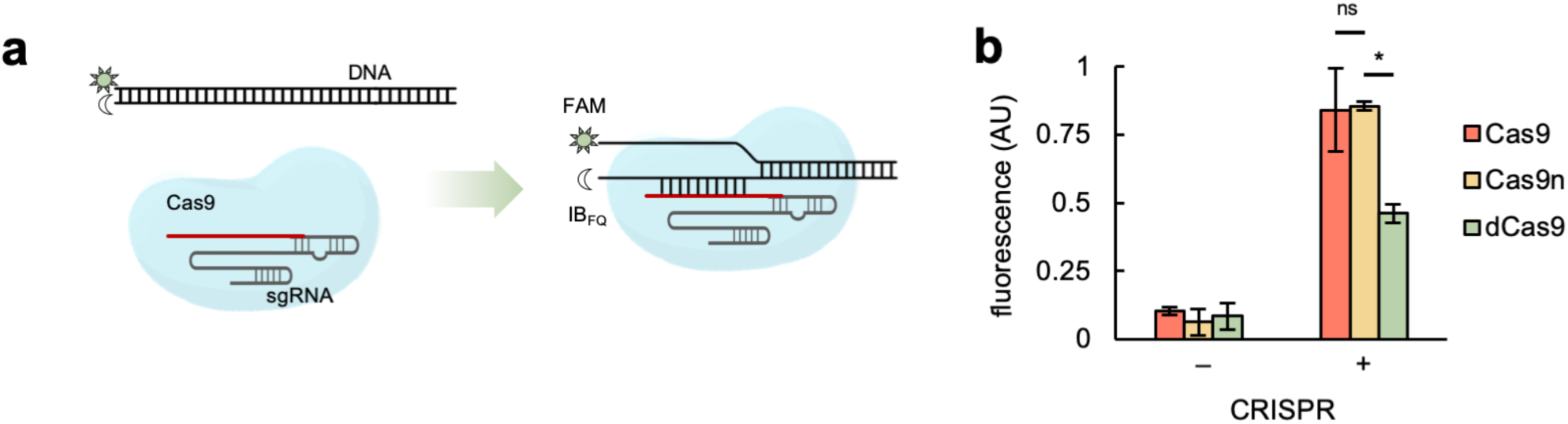
Activity assessment of Cas9 versions. a) Schematics of a test to assess the DNA targeting activity of a CRISPR-Cas9 ribonucleoprotein. The targeted DNA was labelled with a fluorophore (FAM) and quencher (Iowa Black FQ). Upon targeting, the fluorophore is separated from the quencher, then producing a fluorescence signal. b) Fluorescence-based characterization with different Cas9 versions (Cas9, Cas9n, and dCas9). Error bars correspond to standard deviations (*n* = 3). *Statistical significance (Welch’s *t*-test, two-tailed *P* < 0.05). ^ns^Not statistically significant.

**Figure S4:**
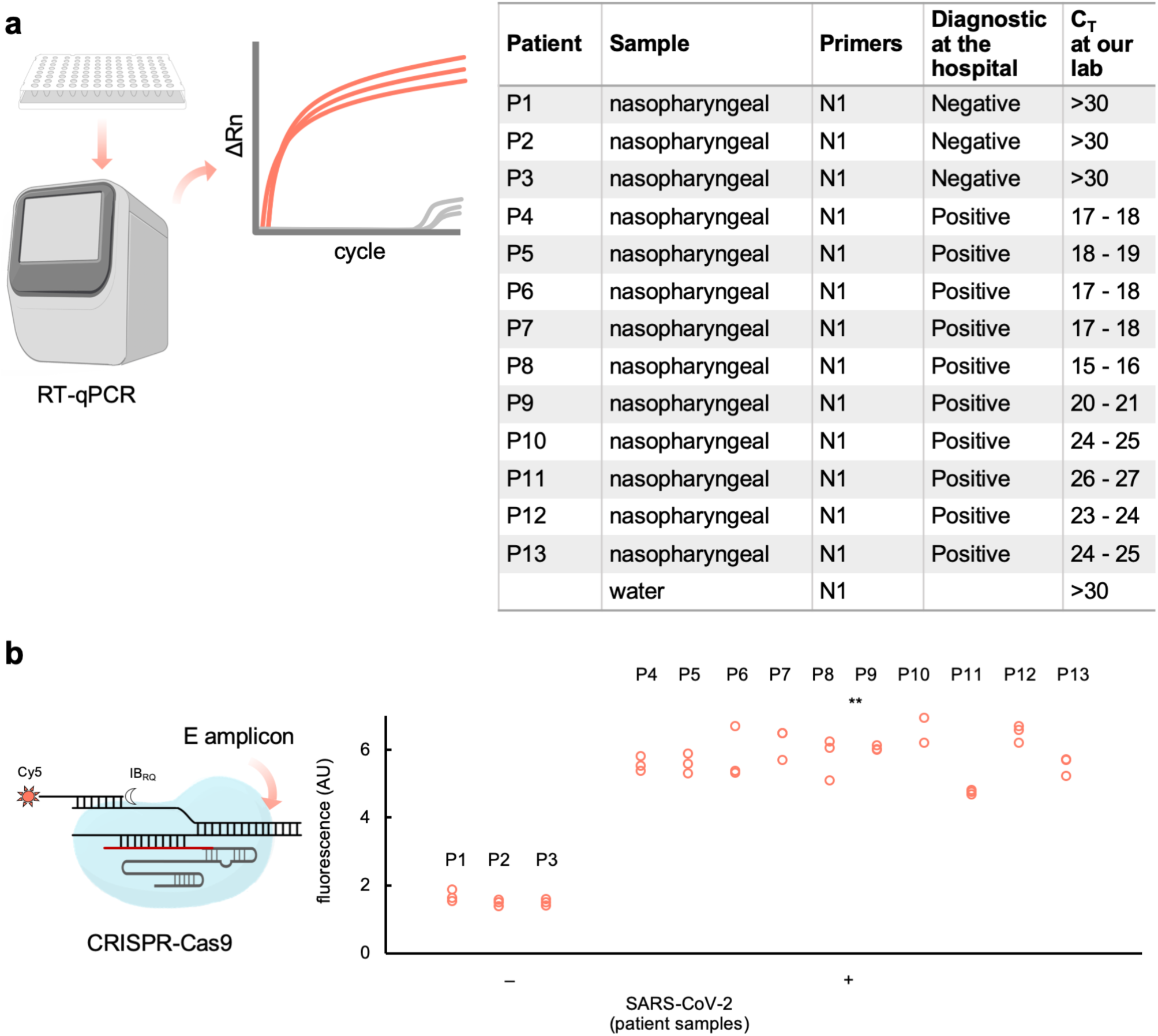
RT-qPCR characterization of patient samples. a) Characteristics of the different patient samples used in this work, including the C_T_ value (N gene). b) Fluorescence-based characterization of all samples with COLUMBO-PCR (E gene). **Overall statistical significance (Welch’s *t*-test, two-tailed *P* < 0.001).

**Figure S5:**
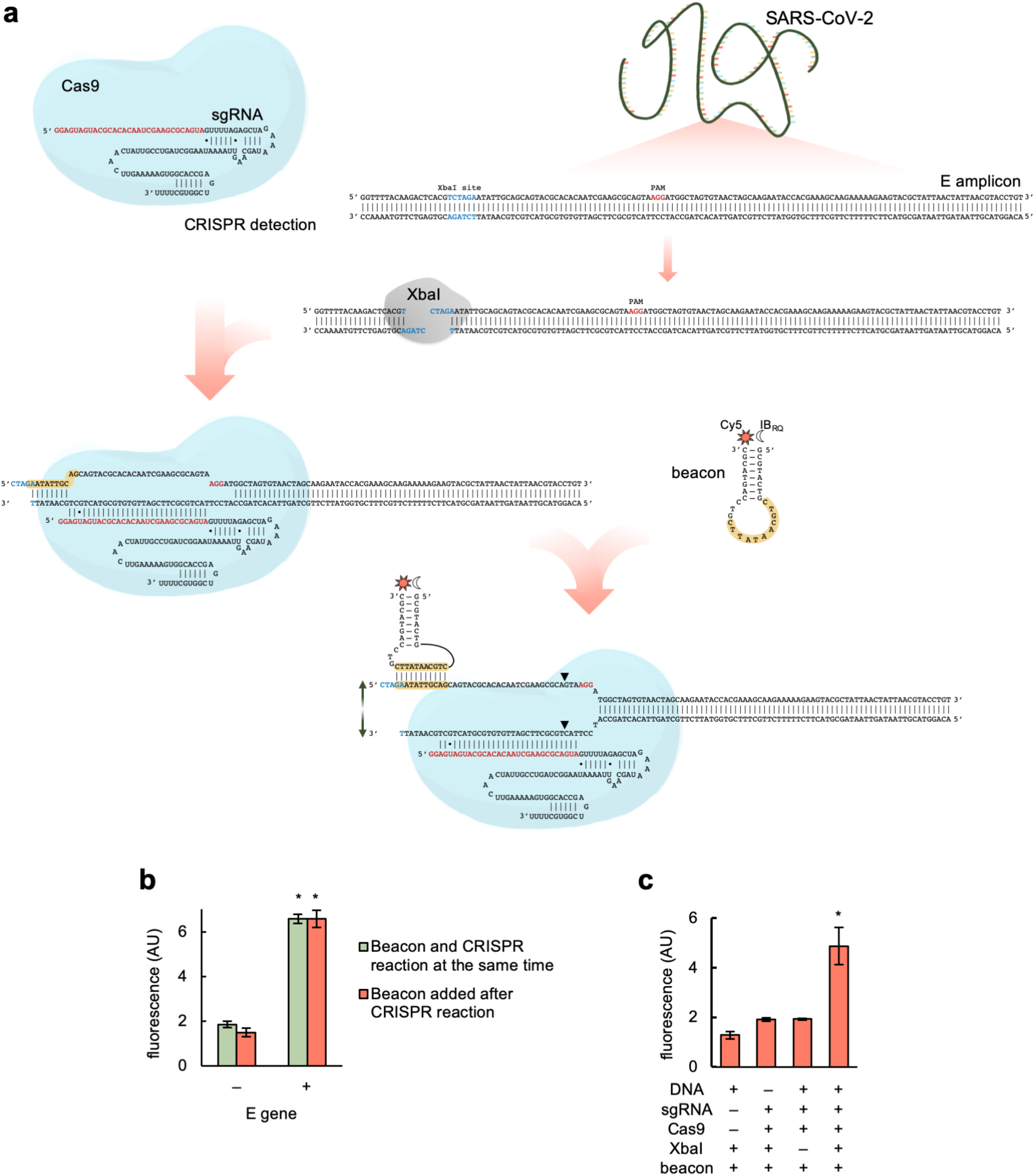
Nucleic acid detection through CRISPR-Cas9-based strand displacement with no prior purification. a) Schematics of the global reaction of amplification and detection of a DNA product from SARS-CoV-2 E gene, containing a PAM (shown in red) for Cas9 recognition and the XbaI restriction site (shown in blue). XbaI was used to cleave the amplicon in the PAM-distal region. A preassembled CRISPR-Cas9 ribonucleoprotein targeting the cleaved amplicon (sgRNA spacer marked in red) was then able to displace a strand so that the molecular beacon could interact with and change its conformation (seed region for this interaction marked in yellow). The molecular beacon was labelled with the fluorophore Cy5 (sun icon) in the 3’ end and a dark quencher (moon icon) in the 5’ end. In this way, the purification step was avoided because the primer and the beacon target different regions. b) Fluorescence-based characterization of the detection when the beacon is added at the beginning of the CRISPR reaction. c) Fluorescence-based characterization of the detection with no prior purification, incubating at the same time the CRISPR-Cas9 ribonucleoprotein, the beacon, and the restriction enzyme (XbaI). Error bars correspond to standard deviations (*n* = 3). *Statistical significance (Welch’s *t*-test, two-tailed *P* < 0.05).

**Figure S6:**
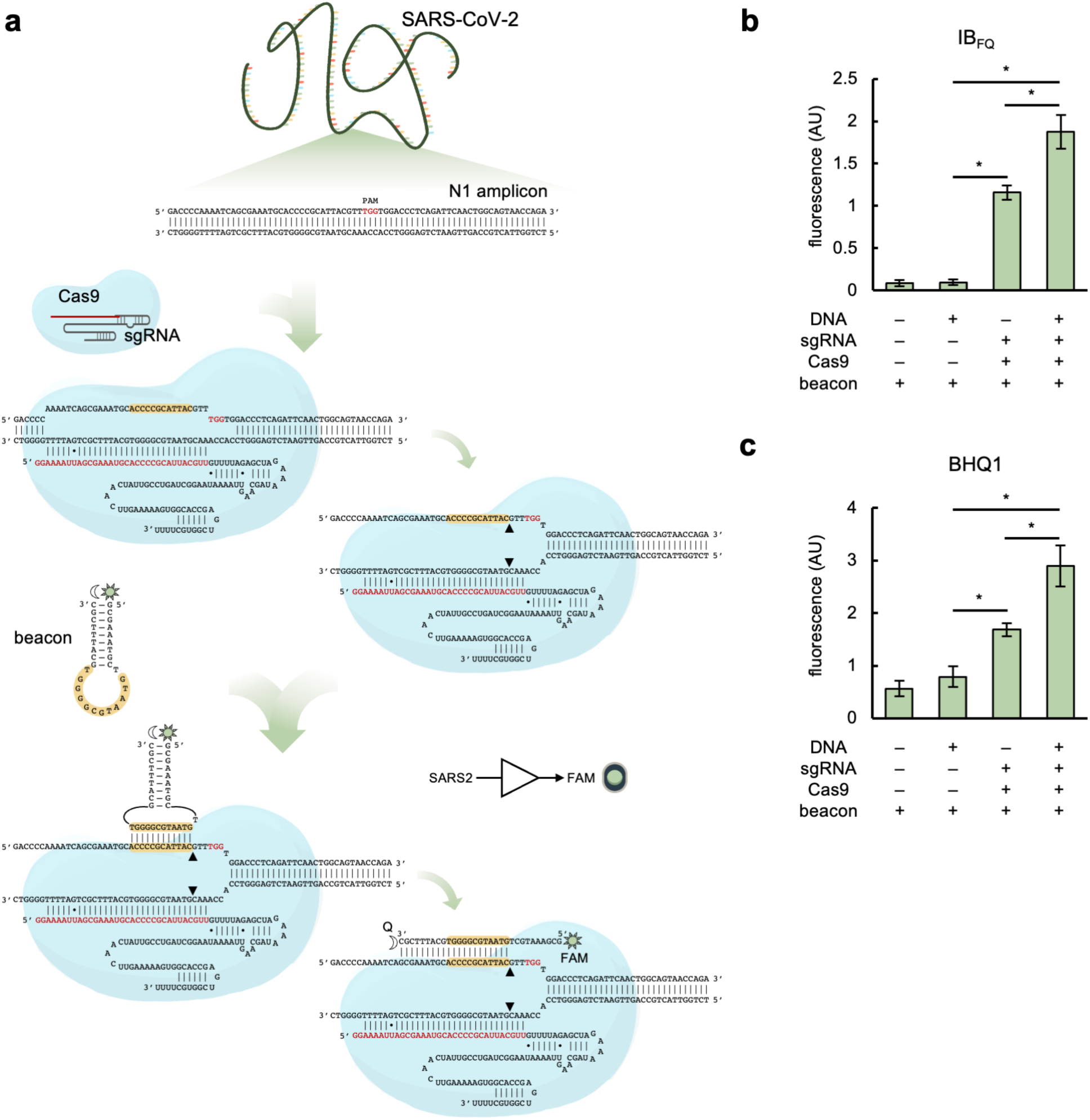
Nucleic acid detection through CRISPR-Cas9-based strand displacement in the PAM-proximal region. a) Schematics of the global reaction of amplification and detection of a DNA product from SARS-CoV-2 N gene (N1 region), containing a PAM (shown in red) for Cas9 recognition. A preassembled CRISPR-Cas9 ribonucleoprotein targeting the amplicon (sgRNA spacer marked in red) was then able to displace a strand so that the molecular beacon could interact with (in the PAM-proximal region) and change its conformation (seed region for this interaction marked in yellow). The molecular beacon was labelled with the fluorophore FAM (sun icon) in the 5’ end and a dark quencher (moon icon) in the 3’ end. b,c) Fluorescence-based characterization of the detection; amplifications performed by PCR. A high signal was observed in absence of the DNA amplicon and presence of the CRISPR-Cas9 ribonucleoprotein, indicating an unwanted interaction between the sgRNA with the beacon (as in this case the sgRNA spacer contains the seed region). In b) the quencher IB_FQ_ was used, while in c) the quencher BHQ1 was used. Error bars correspond to standard deviations (*n* = 3). *Statistical significance (Welch’s *t*-test, two-tailed *P* < 0.05).

**Figure S7:**
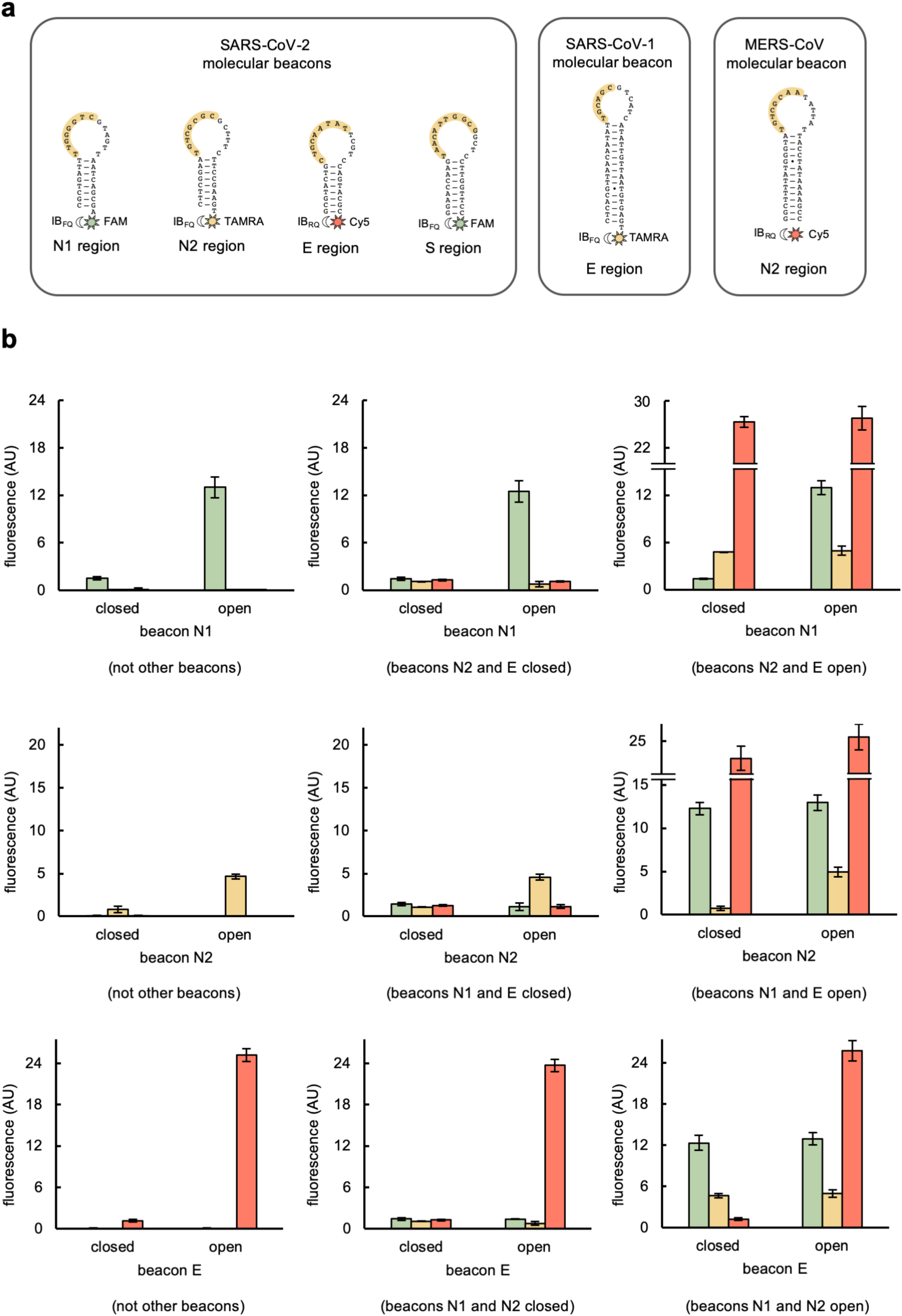
Structure and performance of the molecular beacons. a) Secondary structures of the designed molecular beacons (stem-loop folding) to detect SARS-CoV-2, SARS-CoV-1, and MERS-CoV, labelled with the corresponding fluorophores (sun icons; FAM, TAMRA, or Cy5) in the 3’ end and dark quenchers (moon icons; IB_FQ_ or IB_RQ_) in the 5’ end. The seed region to interact with the displaced strand is marked in yellow. b) Fluorescence-based characterization of the performance of the molecular beacons to detect SARS-CoV-2 when they work alone or in the presence of other beacons. Open beacons obtained by hybridization with appropriate oligonucleotides. Error bars correspond to standard deviations (*n* = 3). Effect of other beacons not statistically significant (one-way ANOVA test, independent samples, *P* > 0.05).

**Figure S8:**
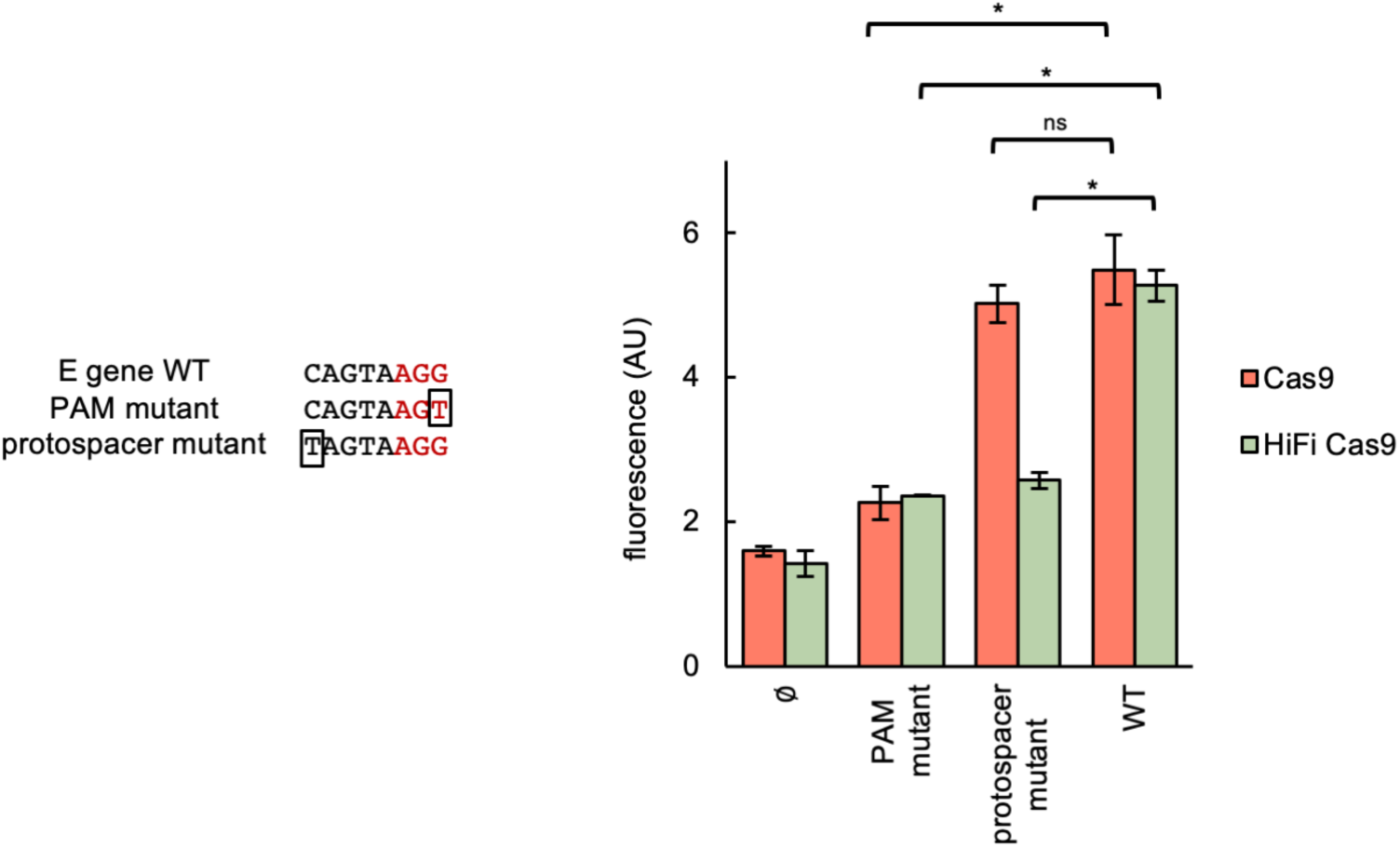
Detection of substitution mutations through CRISPR-Cas9-based strand displacement. Fluorescence-based characterization of E gene variant detection with the native Cas9 and HiFi Cas9. On the left, substitution mutations are framed (PAM shown in red). Error bars correspond to standard deviations (*n* = 3). *Statistical significance (Welch’s *t*-test, two-tailed *P* < 0.05). ^ns^Not statistically significant. These results show that the native Cas9 is only able to discriminate mutations in the PAM, while the HiFi Cas9 is able to discriminate mutations in the PAM and the protospacer, so this latter nuclease seems better suited for applications in which the detection of variants is required.

**Figure S9:**
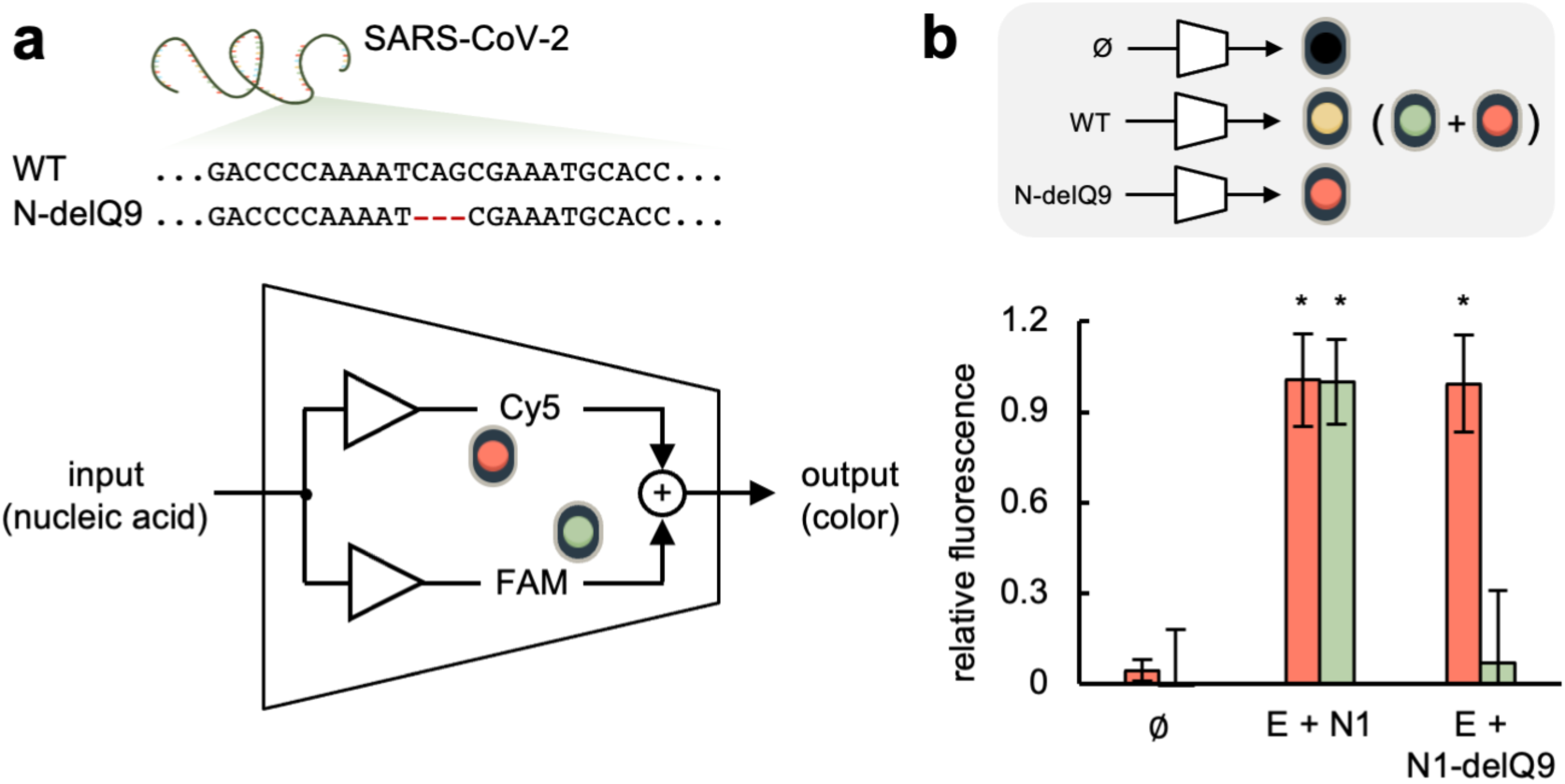
Mutant SARS-CoV-2 detection through CRISPR-Cas9-based strand displacement. a) Schematics of an electronic circuit implementing a molecular program for detection: if the sample is free of SARS-CoV-2, there is no light signal; if it contains the wild-type SARS-CoV-2, a “yellow” signal is obtained (merging the signals from Cy5 and FAM); if it contains a SARS-CoV-2 that carries the mutation N-delQ9, a red signal is obtained (only from Cy5). b) Fluorescence-based characterization of the detection by working directly with DNA amplicons: none, E and N1 (simulating the wild-type SARS-CoV-2), and E and N1-delQ9 (simulating a mutant SARS-CoV-2). Error bars correspond to standard deviations (*n* = 3). *Statistical significance (Welch’s *t*-test, two-tailed *P* < 0.05).

**Figure S10:**
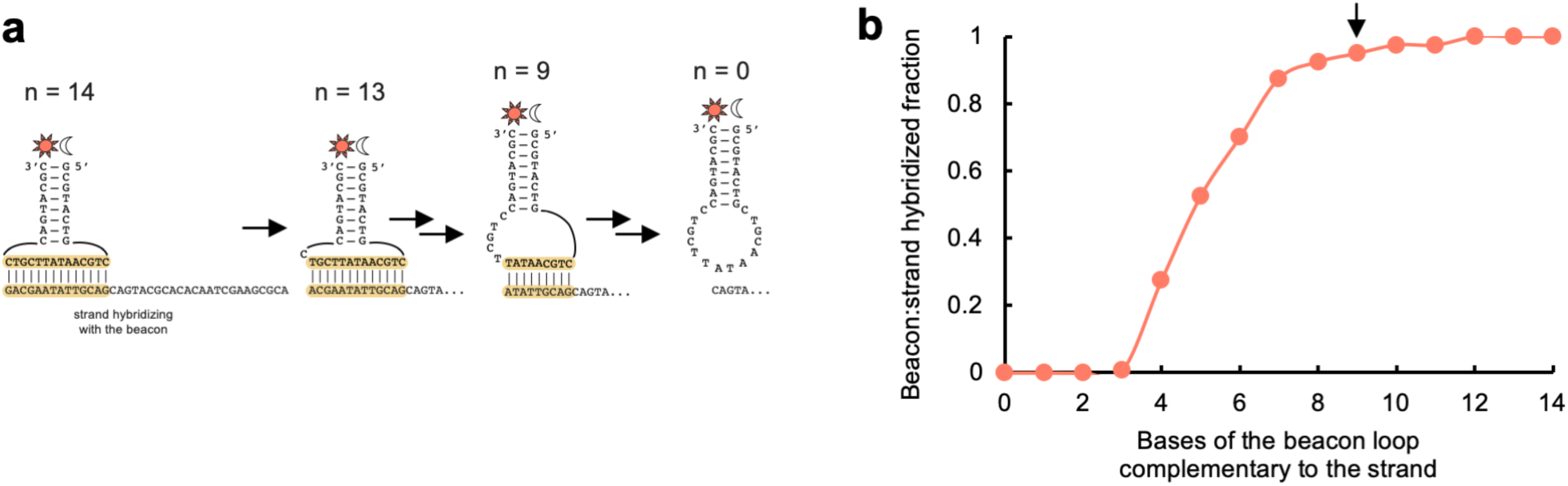
Computational analysis of the energy barrier associated with the interaction of the beacon. a) Schematics of the hybridization between a beacon and a given strand. The beacon for the SARS-CoV-2 E gene was considered (fixed). Different sequences (based on the E gene) were generated to interact with the beacon, starting from one that interacts with the full-length loop (seed region of 14 nt, n = 14) and ending with one that does not interact with the loop (no seed region, n = 0). b) Molar fraction of the hybridized species (beacon:strand) as a function of the size of the seed region (*i.e*., the number of bases of the beacon loop that are complementary to the strand). NUPACK was used considering DNA parameters and a concentration of 40 nM for both species. The arrow marks the sequence corresponding to the actual design.

**Figure S11:**
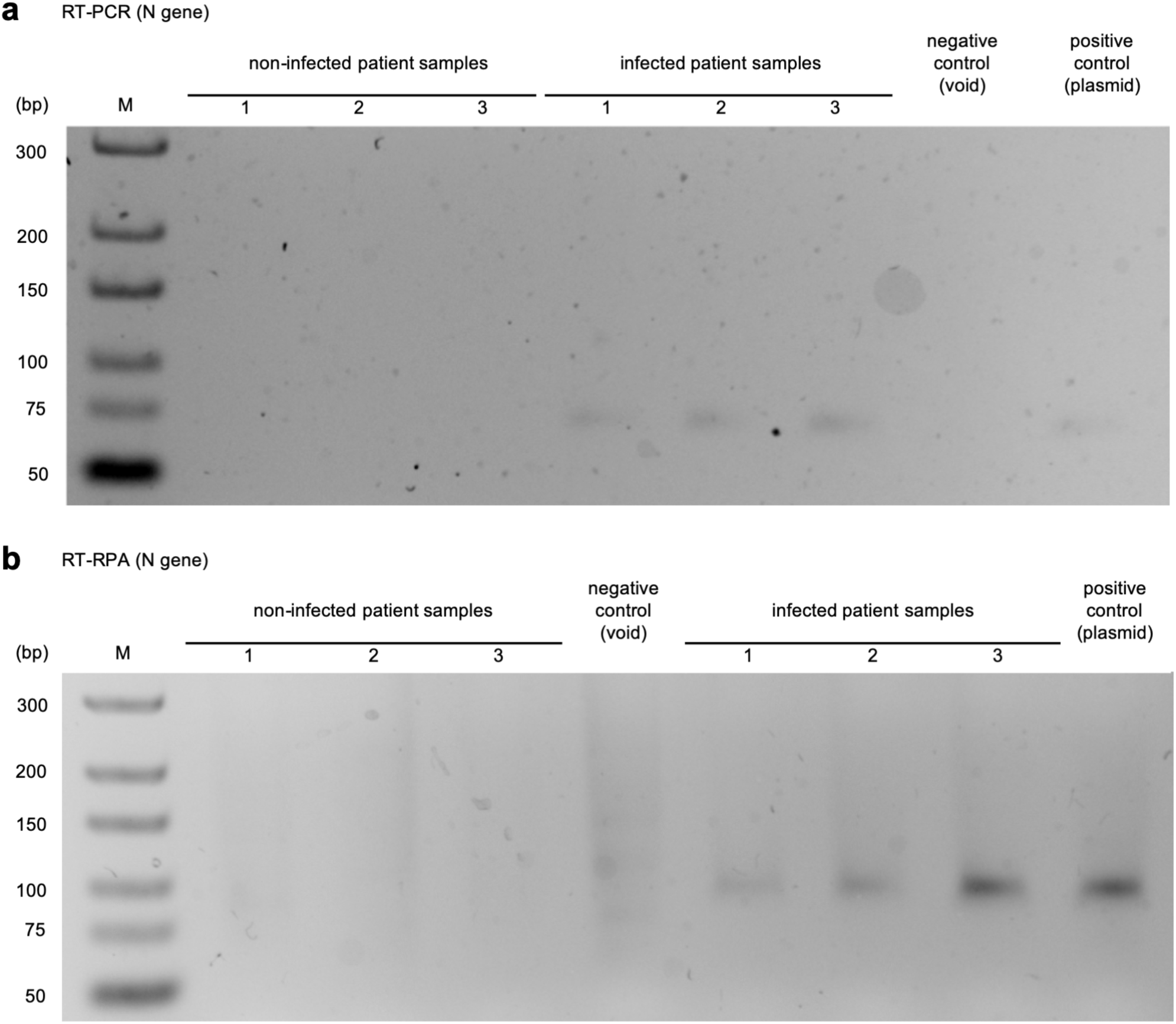
Gel electrophoretic assay to reveal the viral genome amplification. a) Amplification of SARS-CoV-2 (N gene) over patient samples by RT-PCR. b) Amplification of SARS-CoV-2 (N gene) over patient samples by RT-RPA. Due to the RPA buffer composition, we noted that the migration of the fragment in the gel was slower. M, molecular marker (GeneRuler ultra-low range DNA ladder).

## Notes

### Author Declarations

The ethical committee of the Clinic University Hospital approved this study (order #2020/221).

